# Long-term psychological consequences of long Covid: a propensity score matching analysis comparing trajectories of depression and anxiety symptoms before and after contracting long Covid vs short Covid

**DOI:** 10.1101/2022.04.01.22273305

**Authors:** Daisy Fancourt, Andrew Steptoe, Feifei Bu

**Affiliations:** Department of Behavioural Science and Health, university College London, UK

**Keywords:** long Covid, pandemic, mental health, depression, anxiety

## Abstract

**Background:** There is a growing global awareness of the psychological consequences of long Covid, supported by emerging empirical evidence. However, the mergence and long-term trajectories of psychological symptoms following the infection are still unclear.

**Aims:** To examine when psychological symptoms first emerge following the infection with SARS-CoV-2, and the long-term trajectories of psychological symptoms comparing long and short Covid groups.

**Methods:** We analysed longitudinal data from the UCL Covid-19 Social Study (March 2020-November 2021). We included data from adults living in England who reported contracting SARS-CoV-2 by November 2021 (N=3,115). Of these, 15.9% reported having had long Covid (N=495). They were matched to participants who had short Covid using propensity score matching on a variety of demographic, socioeconomic and health covariates (N=962, n=13,325) and data were further analysed using growth curve modelling.

**Results:** Depressive and anxiety symptoms increased immediately following the onset of infection in both long and short Covid groups. But the long Covid group had substantially greater initial increases in depressive symptoms and heightened levels over 22 months follow-up. Initial increases in anxiety were not significantly different between groups, but only the short Covid group experienced an improvement in anxiety over follow-up, leading to widening differences between groups.

**Conclusions:** The findings shed light on the psychobiological pathways involved in the development of psychological symptoms relating to long Covid. The results highlight the need for monitoring of mental health and provision of adequate support to be interwoven with diagnosis and treatment of the physical consequences of long Covid.

## Introduction

Long Covid (defined as the continuation of symptoms that develop during or after acute infection with SARS-CoV-2 that cannot be explained by an alternative diagnosis^12^) is estimated to affect around 10% of people 12 weeks after initial infection, with substantial numbers sustaining symptoms over six months.^1 3^ Long Covid can involve multi-organ complications including affecting the heart, brain, spleen, liver, blood vessels, gastrointestinal tract, kidneys, pancreas, and lungs.^4^ These complications involve multiple overlapping disease mechanisms,^1 9^ and are manifested as a wide range of physiological symptoms (e.g. fatigue, headache, dyspnea, muscle pain, cardiac abnormalities and anosmia) and neurological symptoms (e.g. sleep disturbances, problems concentrating, cognitive impairment).^4 5 6^ However, less well researched are the psychological symptoms associated with long Covid.

Infection with SARS-CoV-2 is, in itself, associated with psychological consequences, including depression, anxiety, stress and adjustment disorders, poorer sleep, increased substance use, and increased use of antidepressants and opioids.^7–10^ Whilst psychological symptoms generally improve over time, some can linger for substantial periods of time (such as 1 year) without much improvement, or can even worsen over time.^12,13^ When focusing specifically on long Covid, a meta-analysis of 39 studies including over 10,000 people found that 19% of people with long Covid reported anxiety and 8% depression as one of their symptoms.^14^ But results from individual studies in some countries have reported much higher prevalence (e.g. 42% for anxiety and 41% for low mood in a UK study)^3^, and anxiety and depression are listed on the UK’s official NHS long COVID symptom list.^15^ However, whilst research is highlighting that psychological symptoms can be a feature of long Covid, a number of questions remain to be answered.

First, when do psychological symptoms of anxiety and depression first emerge in long Covid patients? Mechanistically, the ability of coronaviruses (including SARS-CoV-2) to directly infect the central nervous system (CNS) and cause neuroinflammation and consequential psychiatric symptoms is well reported.^47^ Similarly, SARS-CoV-2 has been found to affect the permeability of the blood-brain barrier, facilitating the entry of peripheral inflammatory cytokines to the CNS, further increasing neuroinflammation.^16,17^ Systemic inflammation as a result of immune-inflammatory dysregulation as a result of SARS-CoV-2 infection has also been shown to contribute to psychiatric and cognitive symptoms in patients.^18^ So it is plausible that heightened anxiety and depression could be experienced soon after SARS-CoV-2 infection. However, increased social isolation due to long Covid symptoms and anxiety related to persistent symptoms could mean that psychological symptoms in fact emerge and increase during and beyond the acute state of infection amongst long Covid patients.^19 20 31^

Second, does the level of psychological symptoms during the acute stage of SARS-CoV-2 infection differ amongst people who will go on to have short Covid vs long Covid? A relationship between the severity of immune-inflammatory dysregulation in patients with SARS-CoV-2 infection and depressive symptoms 3 months later has been reported,^21^ as has a relationship between neuro-immune inflammation and long Covid.^22^ As such, it is possible that initial psychological experiences around the onset of infection could predict an individual’s risk of developing long Covid. But, reciprocally, heightened stress, anxiety and low mood at the onset of illness could itself adversely affect an individual’s recovery from SARS-CoV-2 through increasing or prolonging disruption of neuro-endocrine and neuro-inflammatory processes.^23^ This has been demonstrated both through studies showing bidirectional associations between psychological symptoms and neuro-endo-immune processes as well as genetic studies highlighting pleiotropy between depression and inflammatory processes, suggesting a shared genetic vulnerability that could help explain the relationship between a history of mood disorders and long Covid noted in prior studies.^18^

Third, what are the longer-term trajectories of psychological symptoms amongst people with long Covid beyond the acute stage of infection? To date, much research on this topic has been limited by relatively short-term follow-up (typically of just a few months) and involved limited waves of data collection during that follow-up. Finer-grained data showing trajectories of psychological symptoms over time are still lacking. It is possible that following the acute stage of infection, some of the initial symptoms of psychological distress decline, as can happen for patients with short Covid.^25^ However, it is also plausible that the reverse is true: the process of dealing with debilitating ongoing symptoms could exacerbate psychological distress.

Therefore, this study aimed to explore the mental health trajectories of people experiencing long Covid, compared to those with short Covid using data from the UCL Covid-19 Social Study. To ensure that any potential differences were not due to imbalances between long and short Covid patients in socio-demographic factors, histories of mental and physical health prior to infection, or symptoms of Covid-19 during acute infection, we used propensity score matching (PSM) to construct two balanced groups of long vs short Covid patients. Further, to differentiate experiences post-Covid from usual mental health experiences during the pandemic, we used growth curve models accounting for lockdowns, other social restrictions, and time of the year, tracking individuals in the 10 months prior to their infection with SARS-CoV-2 and for 22 months of follow-up. This study therefore presents one of the largest and longest studies to date of the psychological experiences of patients with long Covid.

## Methods

### Data

Data were derived from the University College London (UCL) Covid-19 Social Study (CSS); a large panel study of the psychological and social experiences of over 75,000 adults (aged 18+) in the UK during the Covid-19 pandemic. The study commenced on 21 March 2020 and involved weekly and then monthly (from August 2020 onwards) online data collection during the pandemic. Participants were recruited via convenience sampling and targeted recruitment of underrepresented/vulnerable groups. The study was approved by the UCL Research Ethics Committee [12467/005] and all participants gave written informed consent. Detailed information on the study is available online at https://osf.io/jm8ra/.

This study used data from participants living in England who completed the special module on Covid-19 experience in November 2021 (N=22 528). Of these, 4,938 participants (22%) reported having had Covid-19 by November 2021. We excluded participants (i) with missing data on Covid-19 specific measures (1%), (ii) who reported infection with SARS-CoV-2 before 21^st^ March 2020 when the study was launched, and (iii) who reported having Covid-19 more than once due to the ambiguity in identifying long Covid-19 dates. This left us with a Covid-19 sample of 3,115 participants (see Supplementary Figure S1 for sample selection).

### Measures

#### Long Covid

In November 2021, the CSS included a special module on Covid-19 experience. Participants were asked ‘overall, do you believe you have ever had Covid-19?’ The response options included: (1) yes, confirmed by a positive Covid-19 test; (2) yes, confirmed by a positive antibody test; (3) yes, suspected by a doctor but not tested; (4) yes, my own suspicions; (5) No, not that I know of. Due to challenges in diagnosis of Covid-19, (e.g. prior to mass testing being available, due to test shortages and access challenges, and due to testing inaccuracies), definitions of long Covid do not require having had a positive test result, but rather the presence of symptoms that were initially suggestive of Covid-19.^26^ So we considered both positive tests and suspected cases (category 1 to 4) as having had Covid-19. The information on date of contract was also collected: ‘If you have had Covid-19, when did you first contract it?’ Moreover, participants were asked if they considered themselves to have or have had long Covid, with responses: (1) yes, a medical professional has formally diagnosed me with long Covid; (2) yes, I have not been formally diagnosed but consider myself to have Long Covid; (3) no, I do not consider myself to have Long Covid; (4) I am unsure. This was recoded into a binary variable (yes/no) with unsure being treated as not having Covid.

#### Mental health outcomes

Depressive symptoms were measured using the Patient Health Questionnaire (PHQ-9);^27^ a standardised instrument for screening for depression in primary care. The questionnaire includes 9 items with 4-point responses with total scores ranging 0-27.

Anxiety symptoms were measured using the Generalized Anxiety Disorder assessment (GAD-7);^28^ a well-validated tool used to screen for generalized anxiety disorder in clinical practice and research. The GAD-7 comprises 7 items with 4-point responses with total scores ranging 0-21.

#### Covariates

To construct comparable groups, a range of factors were taken into account in our analyses. These included gender (women vs men), ethnicity (white vs ethnic minorities), age groups (age 18-29, 30-45, 46-59, 60+), education (up to GCSE levels, A-levels or equivalent, and university degree or above), income (<£16,000, £16,000-29,000, £30,000-59,000, £60-89,000, ≥£90,000 per annum), employment status (employed non-key worker, employed key worker, other), area of living (city, town, rural), living situation (living alone, living with adults only, living with children), number of close friends (0 to 10+) and usual social contacts (twice a month or less, once or twice a week, three times a week or more), self-reported diagnosis of any long-term physical health condition or any disability (yes vs no), and self-reported diagnosis of any long-term mental health condition (yes vs no). Moreover, we included baseline depressive (PHQ-9) and anxiety (GAD-7) symptoms when participants joined the study. Finally, we included a few Covid-19 specific measures, including month when participants had Covid-19 and whether confirmed by a test (Covid-19/antibody test vs doctor’s/own suspicion). In addition, we considered severity of symptoms in the first one to two weeks (severe symptoms vs minor/asymptomatic).

### Statistical Analysis

We employed propensity score matching (PSM) to construct two comparable groups: short Covid-19 versus long Covid-19. The propensity score is the probability of being assigned to a treatment given the value of observed covariates, which is used to pair participants who had long Covid-19 (the treatment group) with those who had short Covid-19 (the control group). The two matched groups should have identical or similar distribution of covariates that are used to estimate the propensity score. We used one-to-one nearest neighbour matching within a calliper (a quarter of one standard deviation of the sample estimated propensity scores). PSM was implemented using Stata psmatch2 command.^29^

To compare mental health growth trajectories between the two matched groups, data were analysed using growth curve models. The time variable was constructed as month since people contracted SARS-CoV-2, which took negative values for the period before infection. This is relative time as the starting point varied across individuals. We controlled for the month (absolute time) in which people contracted SARS-CoV-2 considering an individual’s mental health is subject to the impact of time-related contextual factors (e.g. lockdowns and other restrictions). This was used as a series of dummy variables. Further, the model included a quadratic term to allow for non-linear trajectories (see the Supplement for mathematic equations).

As sensitivity analyses, (i) we reran the analyses using alternative matching methods, namely one-to-many nearest neighbour matching and kernel matching; (ii) we excluded anyone who said they were ‘unsure’ if they had long Covid; and (iii) we explored further people’s understanding of the term ‘long Covid’. If individuals reported believing they had long Covid but subsequently indicated that their symptoms had got better after 2 weeks, they were excluded, as were individuals who reported not having long Covid but who said they had symptoms lasting more than 4 weeks. Analyses were carried out using Stata version 17.

## Results

### Sample Characteristics

Of the 3,115 participants considered for matching, 495 (15.9%) reported having had long Covid-19. 23.8% of them were formally diagnosed by a medical professional and 76.2% self-diagnosed (Table S2). After matching, 481 participants were successfully matched with a control. The propensity score and each individual covariates were balanced after matching (Figure S3 and S4). In total, there were 13,325 observations from 962 participants (13.9 per person).

Sample characteristics by the two matched groups are reported in Table S1. 78.2% of participants in the long Covid-19 group were women. Although women were found to be at a higher risk of long Covid-19^30^, it is more than likely that there was an overrepresentation of women in the sample. However, weights were not applied due to a lack of credible information on the demographic characteristics of people who had long Covid in England. Both groups experienced a comparable range of symptoms from being asymptomatic to being hospitalised (Table S2).

### Growth Trajectories

We initially fitted the growth curve model allowing Long Covid grouping variable to be associated growth rate of the quadratic term. However, there was no evidence supporting this. Therefore, it was removed from the final model.

In the 10 months before contracting Covid-19 there was no evidence that the two Covid-19 groups differed in either the intercept or growth rate of depressive symptoms (Table 1 & Figure 1). However, at the time of infection, people who went on to develop long Covid-19 had 1.3 points higher depressive symptoms on the PHQ-9 (16% higher) than those with short Covid-19. This equated to an increase of 35% in depressive symptoms from the month before they contracted SARS-CoV-2 in the long Covid group vs an increase of 18% in the short Covid group and was independent of all covariates that were included in the PSM model, including initial symptom severity. Over time, there was no evidence that these two groups differed in the growth rate (“Long Covid*time” variable, Table 1), but levels of depressive symptoms remained higher in the Long Covid group (“Long Covid” variable, Table 1) (Figure S5a additionally shows 95% confidence intervals).

**Table 1.**
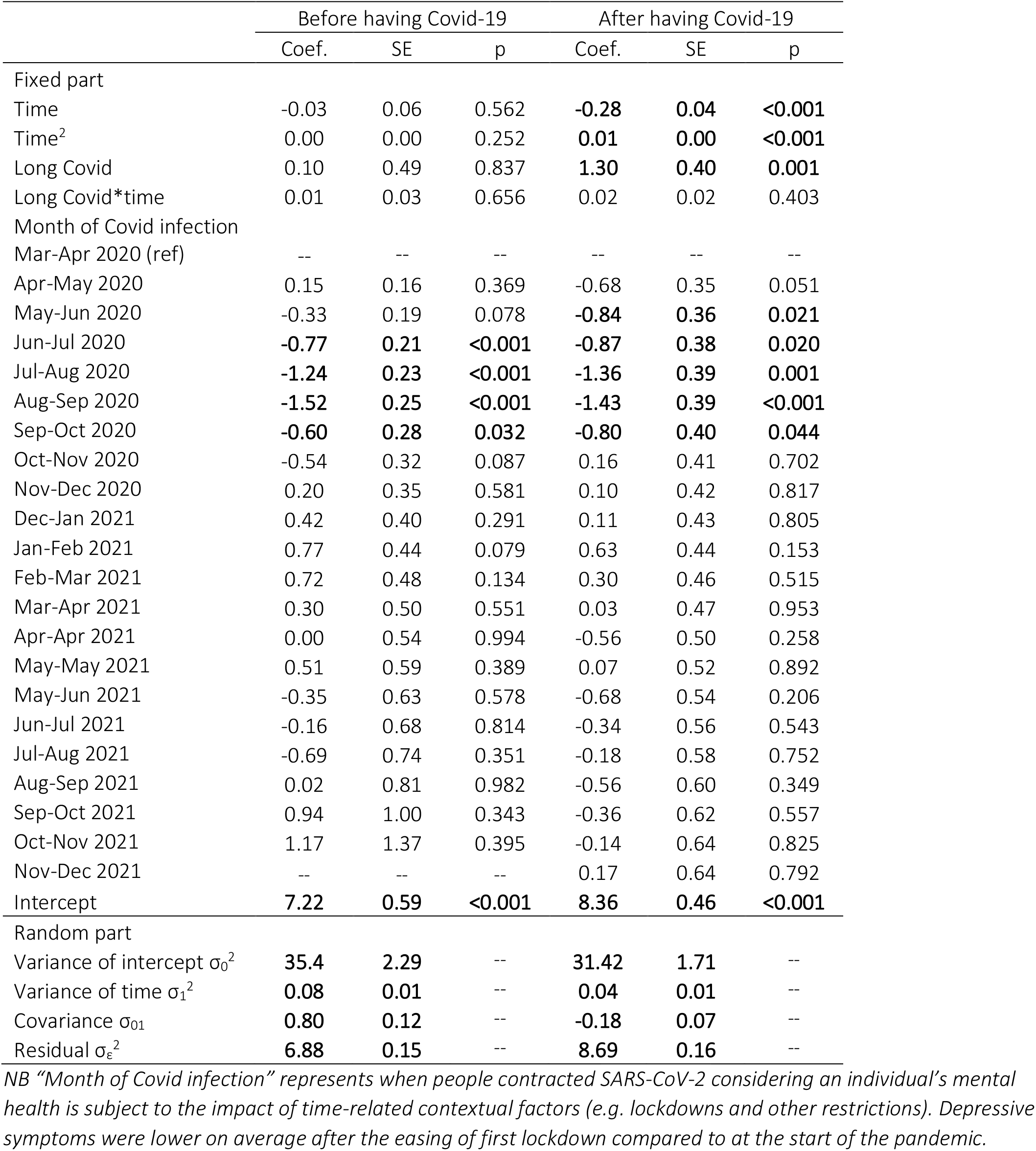
Results from growth curve model on depressive symptoms

**Figure 1.**
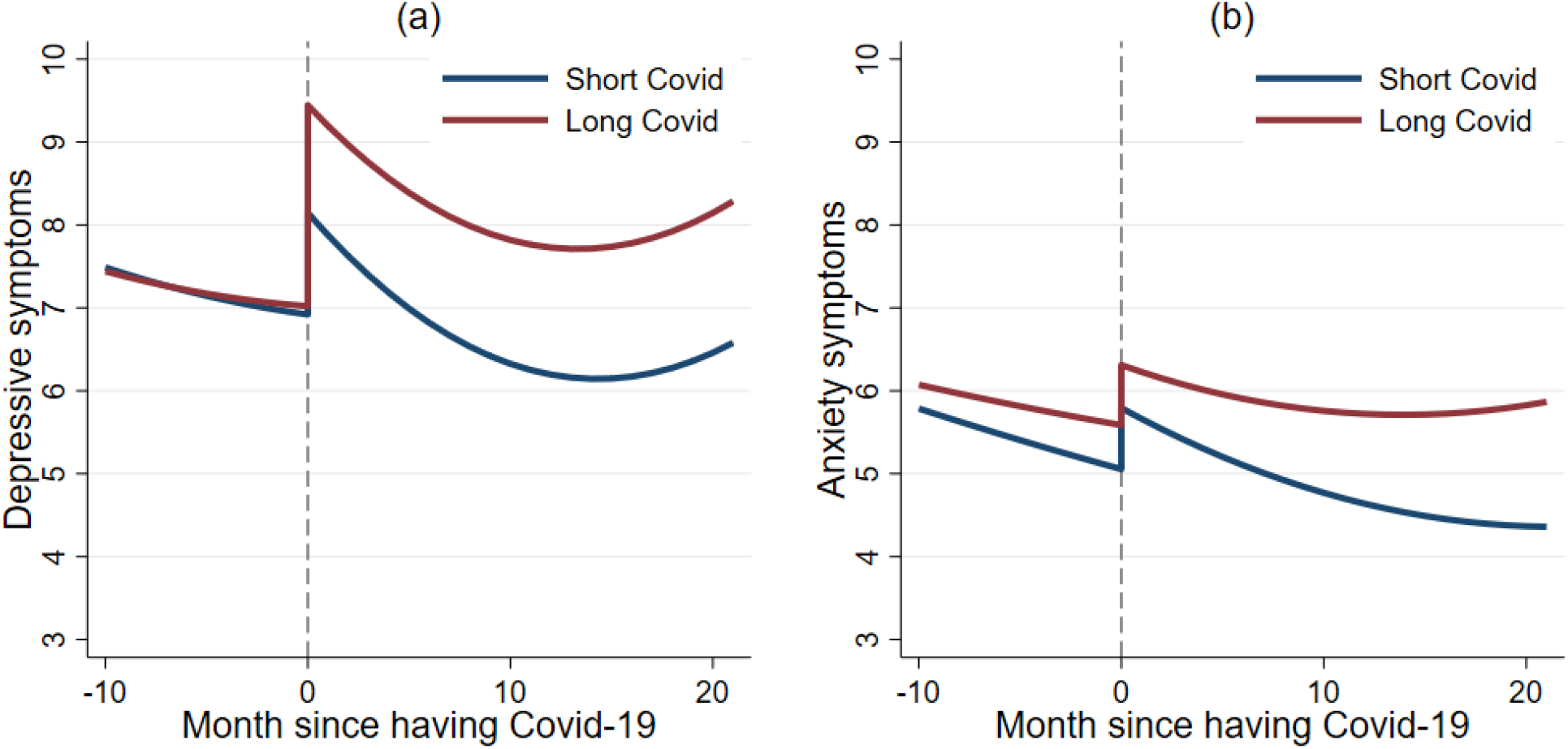
Predicted growth trajectories of depressive (a) and anxiety (b) symptoms from growth curve models

As for anxiety, in the 10 months before contracting Covid-19 there was no evidence that the two Covid-19 groups differed in either the intercept or growth rate of anxiety symptoms (Table 2 & Figure 1). At the time of infection, people who went on to develop long Covid were slightly higher in their anxiety levels to those who had short Covid (0.52 points higher anxiety symptoms on the GAD-7; 8.9% higher), but this was not a significant difference (“Long Covid” variable, Table 2). This equated to an increase of 15% in depressive symptoms from the month before they contracted SARS-CoV-2 in the long Covid group vs an increase of 13% in the short Covid group. However, over time, anxiety symptoms of the long Covid group were less likely to decrease compared with those with short Covid, with an increasing discrepancy in anxiety symptoms by 22 months follow-up (35% higher in the long Covid group) (“Long Covid*time” variable, Table 2) (Figure S5b additionally shows 95% confidence intervals).

**Table 2.**
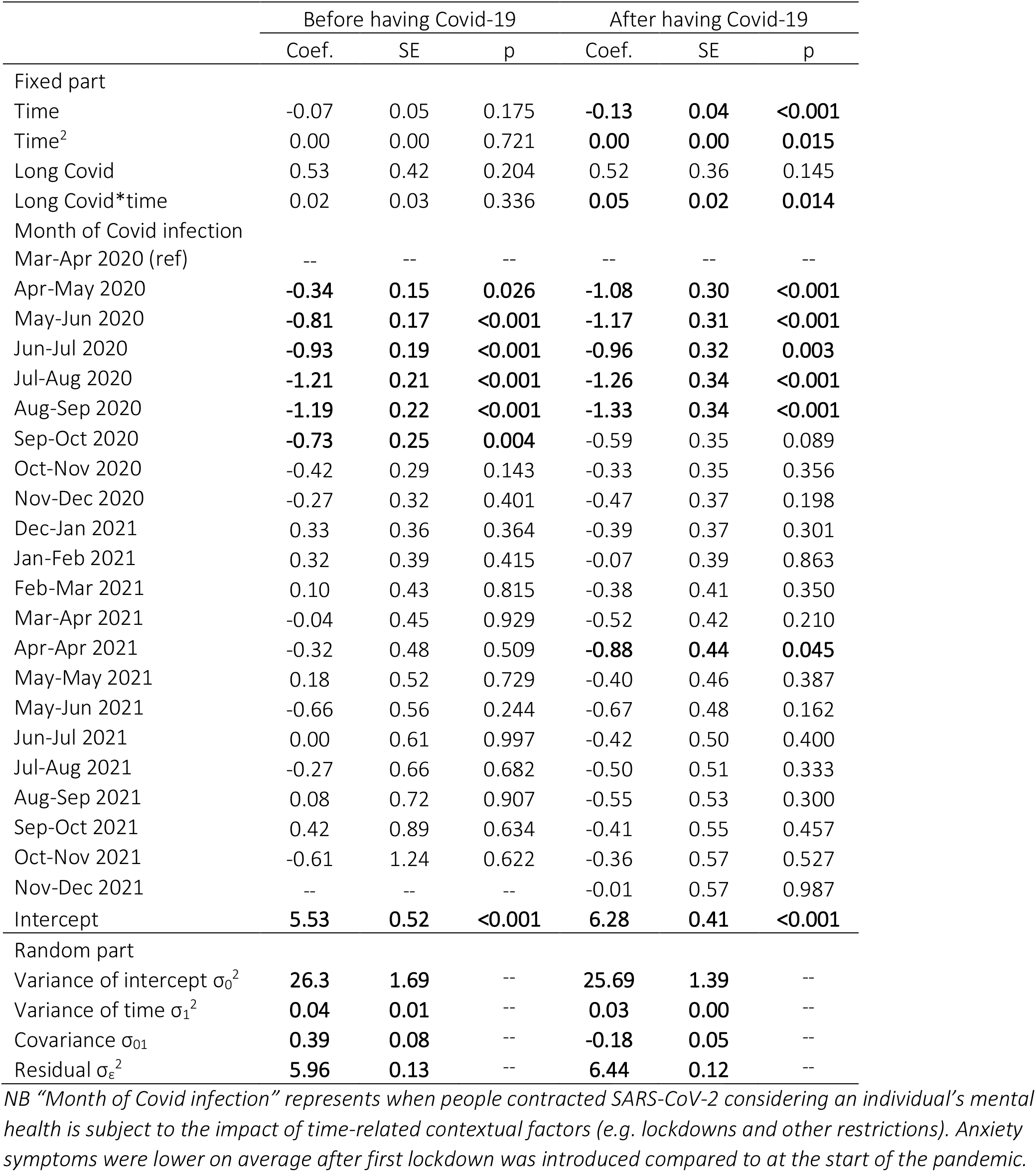
Results from growth curve model on anxiety symptoms

### Sensitivity Analyses

Sensitivity analyses using one-to-many nearest neighbour matching and kernel matching yielded less balanced matches. Further, there was a significant difference in growth factors of depressive and anxiety symptoms between the two matched groups before contracting with Covid-19. The results from the one-to-many nearest neighbour matching are presented in Figure S6 and S7 in the Supplementary Material. Figure S8 shows the predicted growth trajectories by Covid-19 group from kernel matching. Good matches were achieved but results were materially unaffected when applying a more rigorous definition of long Covid (Figure S9) or when excluding people who were unsure about whether they had long Covid or not (Figure S10).

## Discussion

This study examined the mental health trajectories of people who had long Covid-19, compared with those who had short Covid-19. Infection with SARS-CoV-2 was associated with significant increases immediately following the onset of infection in depressive and anxiety symptoms in both groups. In the long Covid group, there was evidence that depression levels rose significantly higher than the short Covid group following onset of Covid-19, but anxiety levels were not significantly different between groups. Over time, the long Covid group maintained heightened levels of depression symptoms compared to the short Covid group, and for anxiety the long Covid group did not experience the improvements in anxiety seen in the short Covid group, leading to widening differences in mental distress between the two groups.

This study sheds light on the mechanisms at play in the development of long Covid. First, symptoms of depression and anxiety emerge quickly following the onset of SARS-CoV-2 infection, with the peak in both symptoms occurring at the first measurement timepoint post-infection (i.e. within 1 week of reported infection). This points to immediate psychobiological pathways being implicated in the aetiology of Covid-related mental distress both in long and short Covid. Existing literature proposes that inflammatory mechanisms could be at play,^47^ but it is also possible that fear related to contracting the virus could also exacerbate mental distress, especially since levels of worries about catching SARS-CoV-2 and potentially becoming seriously ill from it have been associated with heightened anxiety and depression across the pandemic.^31^ Whilst our study included participants who had a range of symptom experiences from hospitalisation to symptoms being mild or even asymptomatic, all participants in the study either knew (via testing) or suspected that they had SARS-CoV-2. We were unable to ascertain whether the reported increases in anxiety and depression found here were primarily driven by biological processes or psychological processes (i.e. fear of the virus) as we lacked data on psychological experiences in people with SARS-CoV-2 who were asymptomatic and did not know they had the virus.

Second, even when long and short Covid patients were matched on physical and psychiatric comorbidities prior to Covid and levels of SARS-CoV-2 symptom severity and even though they displayed similar trajectories of anxiety and depressive symptoms in the 10 months prior to infection, those in the long Covid group experienced greater increases in depressive symptoms immediately following the onset of SARS-CoV-2 infection. This complements previous work suggesting that the ability of SARS-CoV-2 to raise levels of systemic and neuro-inflammation may be greater in patients with long Covid and also suggests that whether an individual is likely to experience long Covid is decided early on in their experience of the virus.^21 22^ Notably, there was not a significant difference in initial increases in anxiety levels between the two groups, which would support theories that the increase in depressive symptoms amongst long Covid patients is biologically driven by the virus rather than a manifestation of a greater predisposition to emotional distress during illness.

However, despite the larger initial increase in depressive symptoms in long Covid patients, both long Covid and short Covid groups experienced parallel decreases in symptoms over time (even within one month of contracting SARS-CoV-2, despite long Covid patients citing their physical symptoms persisting beyond this). This could suggest a process of psychological recovery, even potentially whilst experiencing ongoing physiological symptoms. However, we did not ask participants about how their symptoms and functioning at follow-up compared to when their virus started. So some of this improvement in depressive symptoms amongst long Covid patients could be illustrative of an improvement in long Covid itself (potentially mirroring reductions in levels of inflammation), whilst for patients who continued to experience physiological symptoms, this psychological recovery may not have been felt. Further, despite some improvement, levels of depressive symptoms remained higher in the long Covid group across the entire 22-month follow-up than in the 10 months prior to infection, whereas the short Covid group returned to baseline or below baseline levels within 4 months. So any apparent recovery was not complete. Relatedly, levels of anxiety did not improve over time in long Covid patients over the follow-up, with an increasing discrepancy compared to short Covid patients. The evident persistence of mental distress in long Covid patients mirrors findings from previous studies of patients experiencing other coronaviruses. For example, patients who developed SARS-CoV (“SARS”) in 2003-2004 reported persistent stress one year later without signs of decrease, even if physical symptoms had improved.^32,33^ But it may not simply be a case of initial anxiety symptoms experienced during acute infection (whether initially biologically or psychologically driven) persisting, but also a consequence of new psychological challenges relating to the realisation that one’s initial infection is becoming long Covid and the associated psychological, social and behavioural consequences of ongoing illness (e.g. challenges accessing treatments, threat’s to one’s identity and uncertainty about the future). ^19 20 31^ Notably, whilst there was a slight increase in depressive symptoms in both groups towards the end of the follow-up, the confidence intervals widened across the follow-up period as numbers of people who had contracted SARS-CoV-2 early enough in the pandemic to be followed-up 22 months later decreased. So this increase is likely a feature of the sample rather than a common pattern amongst both groups over time.

Our findings highlight that for patients with long Covid, psychological symptoms could persist for as long as 2 years post-infection, with clear consequences the treatment of long Covid patients. Barriers to diagnosis with long Covid and subsequent challenges navigating and accessing treatments can in itself exacerbate long Covid symptoms.^20^ Given the well-established interconnection between psychological and inflammatory processes, these additional treatment-related stressors could contribute to the prolonging of long Covid symptoms. So it is important that patients feel listened to, validated, and supported in their diagnosis and treatment.^20^ There has been a call for long Covid public health response to include personalised treatment and rehabilitation and multidisciplinary care.^36^ Our findings support this call, and further suggest that (i) clinicians should be aware of the possibility for depression, anxiety and other psychiatric symptoms in patients with long Covid and recognise that such symptoms may not just be a consequence of ongoing physical symptoms but may have been primary outcomes from the initial SARS-CoV-2 infection; (ii) neuropsychological evaluations for patients who are experiencing long Covid could be valuable to determine whether specialised mental health support is also required, and such evaluations should not assume a resolution of psychological symptoms in the absence of treatment; and (iii) psychological support should be provided as part of multidisciplinary care and, given the early onset of psychological symptoms, early provision of such for patients suspected of having long Covid should be recommended.^7^ In some countries such as the UK, it has been recommended that mental health problems alongside or as a result of long Covid can be managed by following existing relevant guidelines.^4^ However, we recommend that any mental health support needs to be provided alongside (rather than as a substitute for) broader medical investigation and support for long Covid, given diagnosis of psychiatric symptoms without adequate attention to other symptoms has been found to be detrimental to mental health in patients with long Covid.^37^ Finally, informal mental health support for patients experiencing long Covid such as peer support groups and community social and cultural activities via schemes such as social prescribing should be encouraged as evidence is already suggesting that the broader support and validation of experiences from others can assist in recovery.^20^

A main strength of this study lies in repeated monthly follow-ups of the same participant over 22 months since March 2020, allowing the longest and most detailed follow-up to date on psychological experiences of long Covid patients. By using propensity score matching, we were able to simulate a randomised trial with the measured covariates randomised between people who had long Covid and those who had regular Covid. We achieved a good quality match, which resulted in equivalent mental health scores in the months prior to infection, improving our ability to infer that any subsequent changes in mental health from immediately after infection were as a result of that infection. However, we cannot rule out the possibility of potential biases due to unobserved covariates that are associated with the risk of having long COVID-19 (although given the wide range of covariates included in our models, any remaining unobserved heterogeneity should be relatively small). Moreover, whilst our sample showed good heterogeneity in initial severity of SARS-CoV-2 symptoms and subsequent long Covid experiences, our sample may not be representative of the population who had (long) Covid, especially as those with more severe ongoing long Covid symptoms may have been more likely to drop out of the study. We followed current best practice in epidemiological research in how we defined long Covid. But some of our patients lacked formal diagnoses, so it is possible that some participants’ symptoms were caused by alternative illnesses. Additionally, our sample was relatively heterogeneous in terms of long Covid experiences, with a range of different patterns of symptoms reported, from persistent acute symptoms for months, to relapsing-remitting symptoms, to low-level persistent symptoms. Whilst this heterogeneity is typical in the diagnosis category of “long Covid”, future research could consider whether there are differential psychological experiences depending on pattern of long Covid symptoms. Finally, our sample relied on participants’ self-report of the date they contracted SARS-CoV-2. This could have been affected by recall bias. So whilst we found an increase in psychological symptoms immediately following reported onset of infection, this increase may have lagged by a few days. Nonetheless, this relatively short lag time does not undermine the conclusions or proposed mechanisms.

Overall, this study presents the longest and most detailed data on psychological experiences of long Covid patients to date, answering crucial questions. Psychological symptoms of anxiety and depression emerge quickly following onset of SARS-CoV-2 in long Covid patients, with levels of depression rising significantly above levels in patients who go on to experience short Covid. Over the following two years, whilst there is some decrease in depressive symptoms, they remain above pre-infection levels, whilst anxiety levels show little signs of improvement. This presents a contrast to short Covid patients where levels return to below-infection levels within 4 months of infection. The findings shed light on the psychobiological pathways involved in the development of psychological symptoms relating to long Covid as well as suggesting that initial psychological experiences during SARS-CoV-2 infection could be explored further as potential predictors of subsequent risk of developing long Covid. The results also show that we cannot assume a gradual organic recovery from the psychological effects of long Covid, highlighting the need for monitoring of mental health and provision of adequate support to be interwoven with diagnosis and treatment of the physical consequences of long Covid.

## Data Availability

Anonymous data will be made publicly available following the end of the pandemic.

## Declarations

### Ethics approval and consent to participate

The Covid-19 Social Study was approved by the UCL Research Ethics Committee [12467/005] and all participants gave written informed consent.

### Availability of data and materials

Anonymous data will be made publicly available following the end of the pandemic.

### Author contributions

DF and AS developed the study idea. FB analysed the data. DF and FB wrote the first draft. All authors had accessed and verified the underlying data. All authors provided critical revisions, read and approved the submitted manuscript.

## Acknowledgements

The researchers are grateful for the support of a number of organisations with their recruitment efforts including: the UKRI Mental Health Networks, Find Out Now, UCL BioResource, SEO Works, FieldworkHub, and Optimal Workshop. The study was also supported by HealthWise Wales, the Health and Care Research Wales initiative, which is led by Cardiff University in collaboration with SAIL, Swansea University.

