## Supplement for "Long-term psychological consequences of long Covid: a propensity score matching analysis comparing trajectories of depression and anxiety symptoms before and after contracting long Covid vs short Covid"

### Supplementary Material

Model specification:

$$Y_{ij} = \pi_{0i} + \pi_{1i}time_{ij} \; + \; \pi_{2i}time_{ij}^2 \; + \varepsilon_{ij} \qquad \text{(Eq. 1)}$$

$$\pi_{0i} = \gamma_{00} + \gamma_{01}group_i + \gamma_{01}month_i + \zeta_{0i} \qquad \text{(Eq. 2)}$$

$$\pi_{1i} = \gamma_{10} + \gamma_{11}group_i + \zeta_{1i} \qquad \text{(Eq. 3)}$$

$$\pi_{2i} = \gamma_{20} + \gamma_{21}group_i + \zeta_{2i} \qquad \text{(Eq. 4)}$$

The growth curve model is presented mathematically in Equation (1) to (4). Equation (1) addresses intra-individual growth where $Y_{ij}$, depressive or anxiety symptoms for the individual i at time j, is a function of time and its quadratic term. $\pi_{0i}$ represents the individual i's initial status. $\pi_{1i}$ and $\pi_{2i}$ represent growth rates and $\varepsilon_{ij}$ is the residual term. Equation (2) to (4) addresses inter-individual differences in the initial status (intercept) and growth rates. $\gamma_{00}$, $\gamma_{10}$and $\gamma_{20}$ represent the population average initial status (intercept) and growth rates. $\zeta_{0i}$, $\zeta_{1i}$ and $\zeta_{2i}$ are parameter residuals.

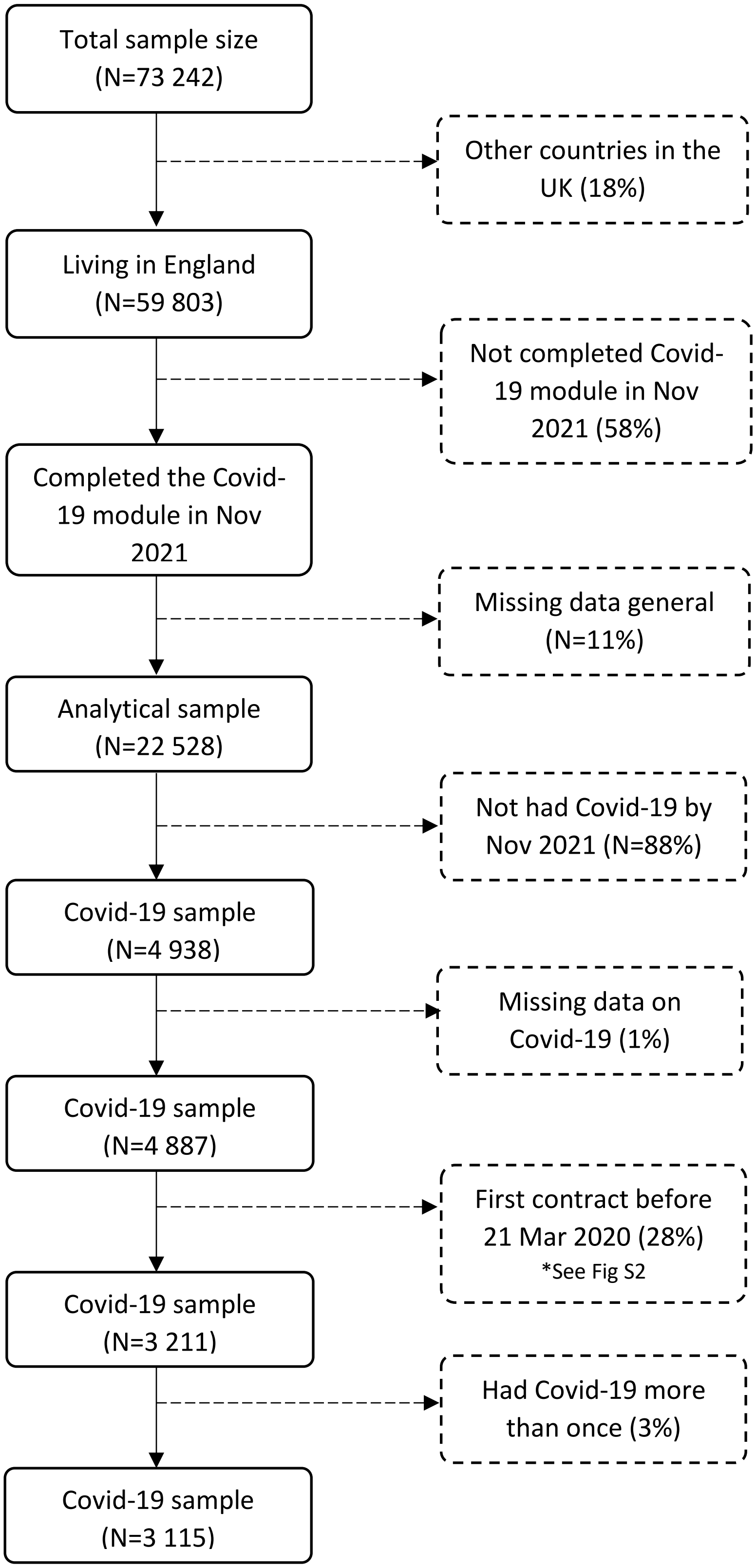


Fig S1. Sample selection process

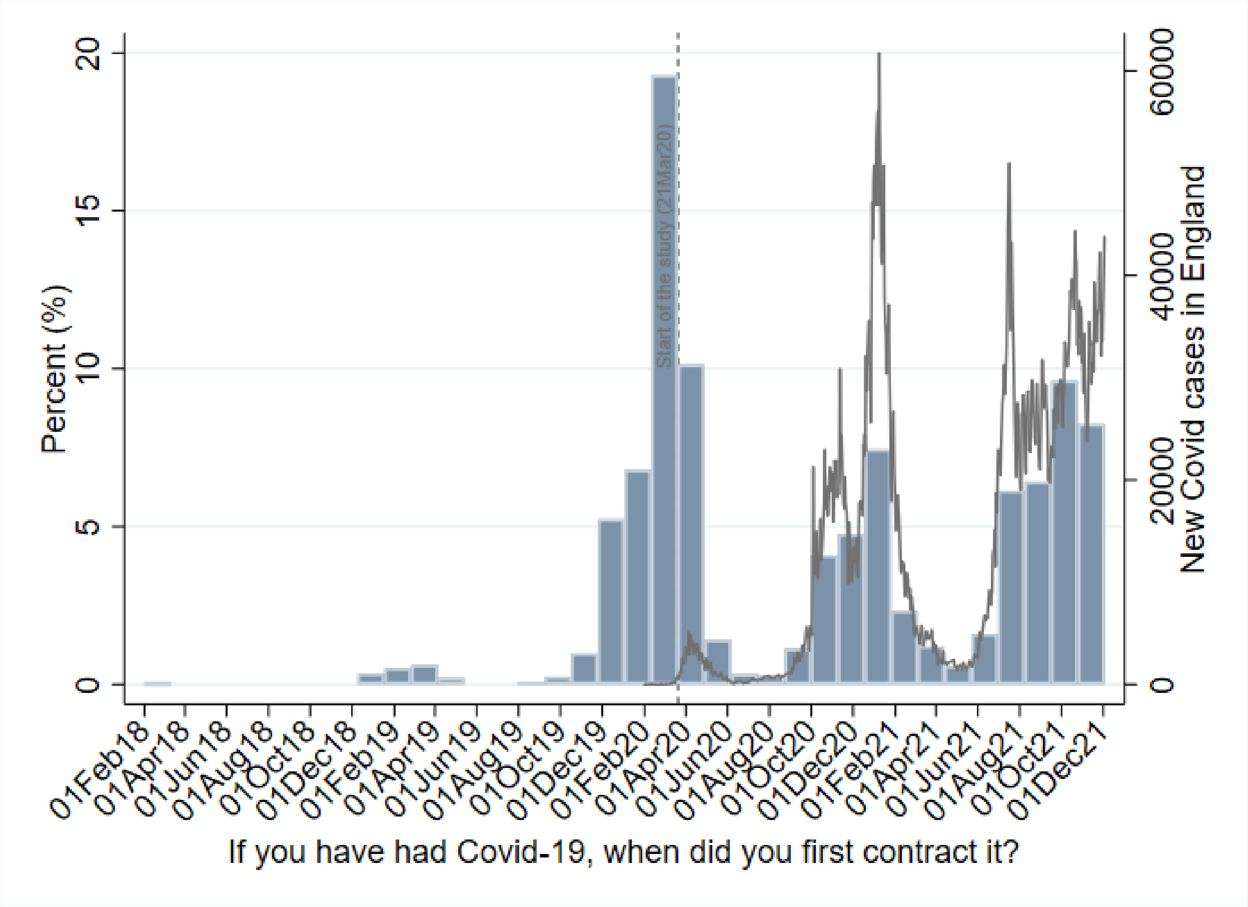


Fig S2. The date of first contract (N=4,938)

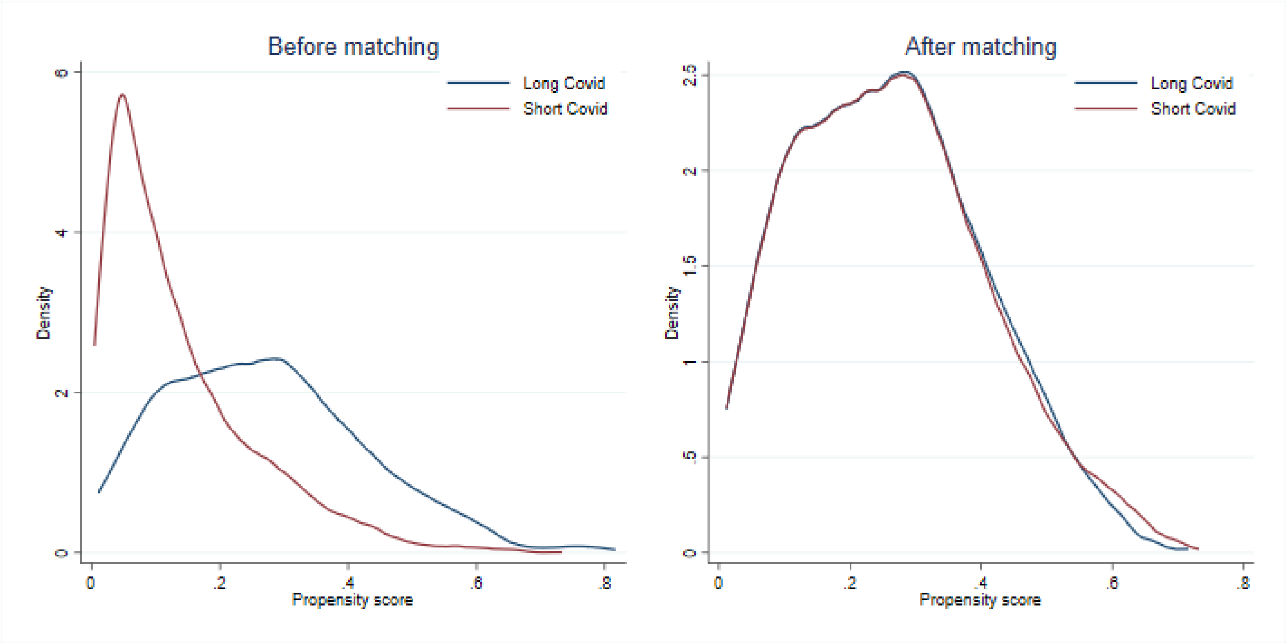


Fig S3. Propensity score density before and after matching

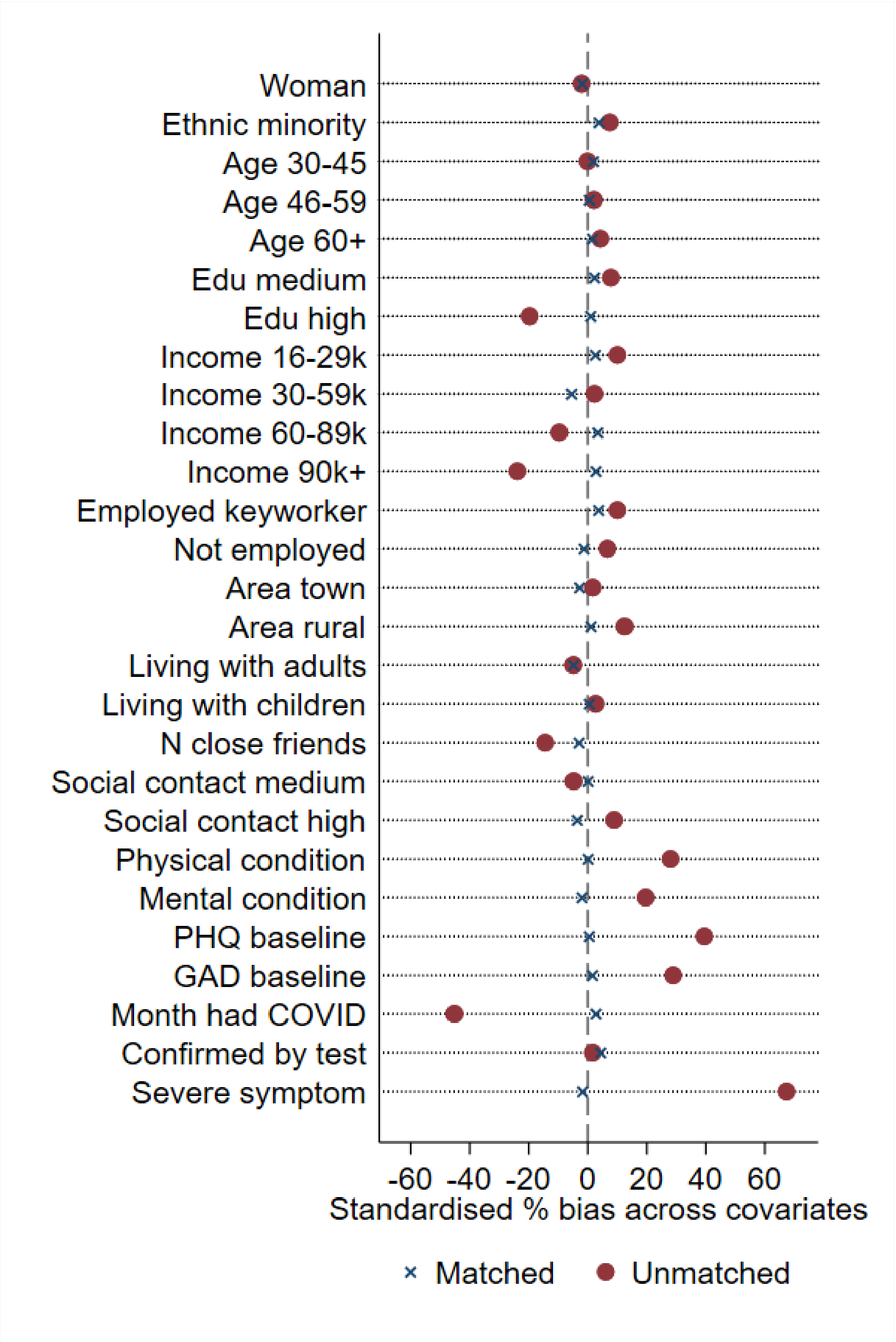


Fig S4. Standardised percentage bias for each covariate before and after matching

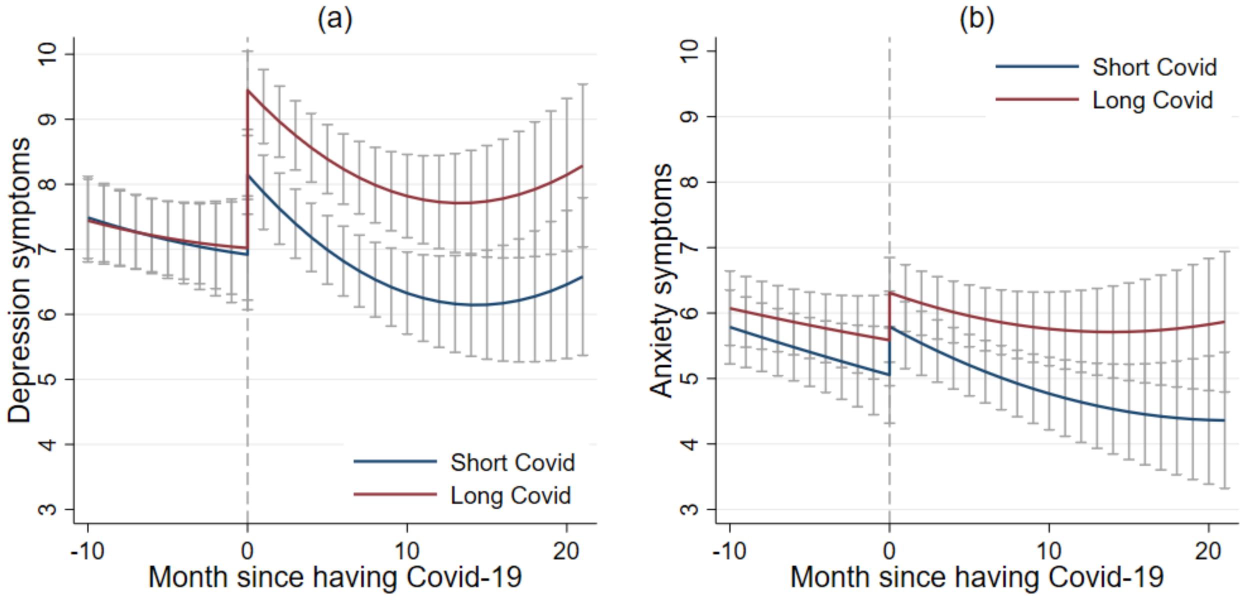


Fig S5. Predicted growth trajectories by Covid-19 group with 95% confidence intervals

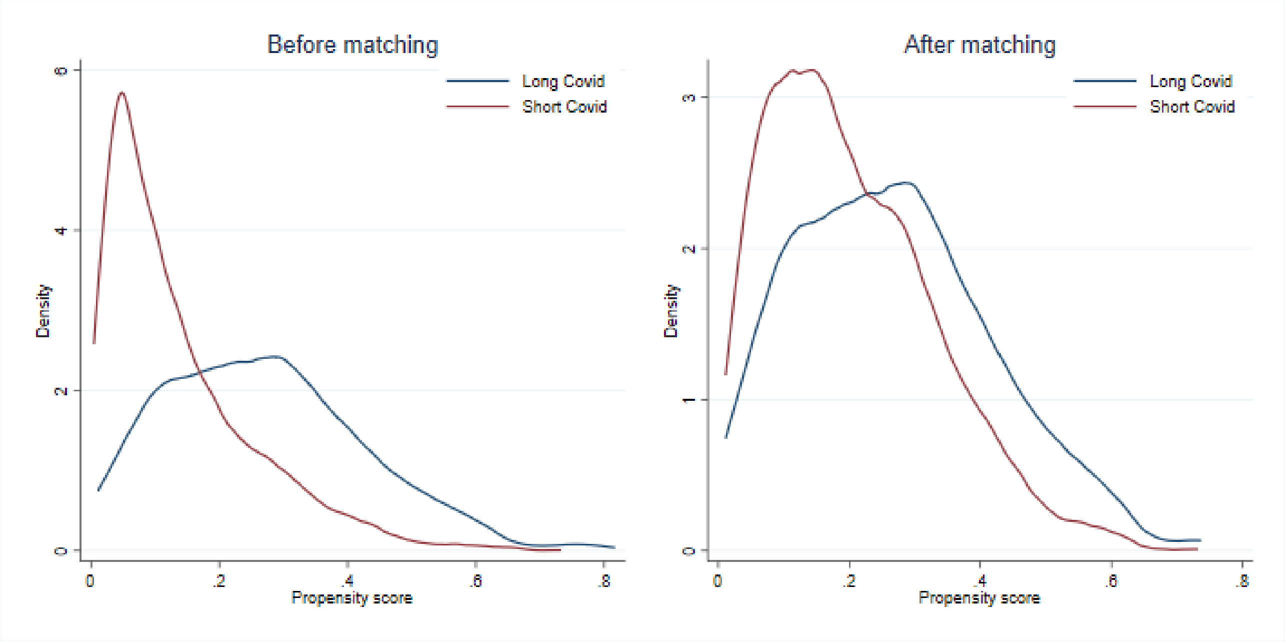


Fig S6. Propensity score density before and after matching based on one-to-many nearest neighbour matching

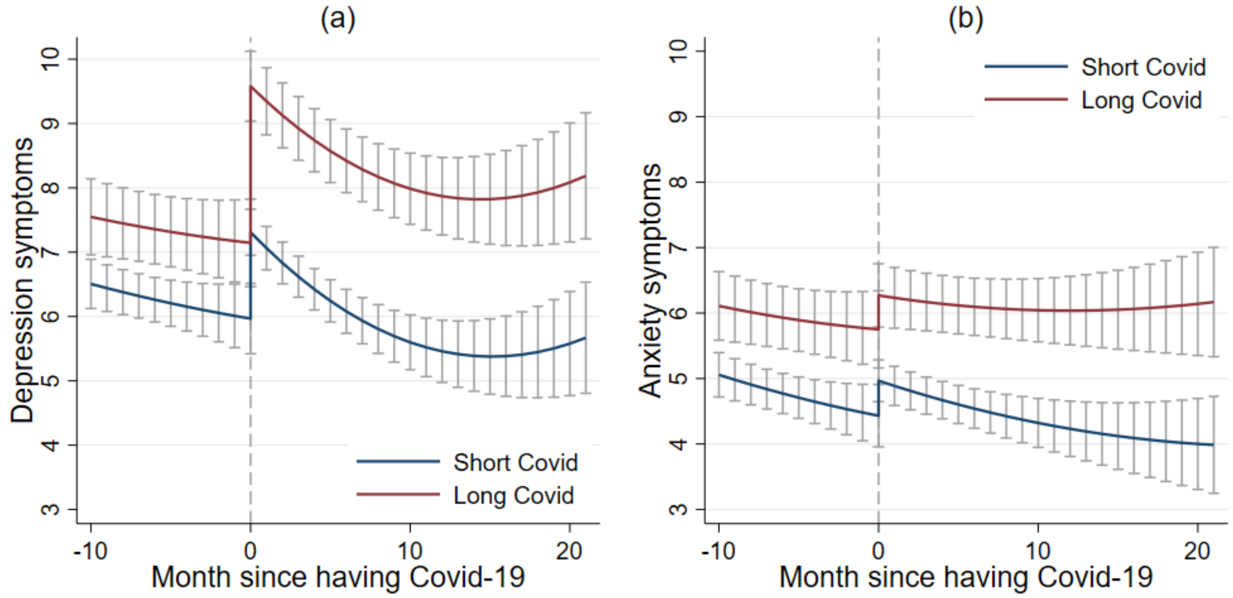


Fig S7. Predicted growth trajectories by Covid-19 group with 95% confidence intervals based on one-to-many nearest neighbour matching

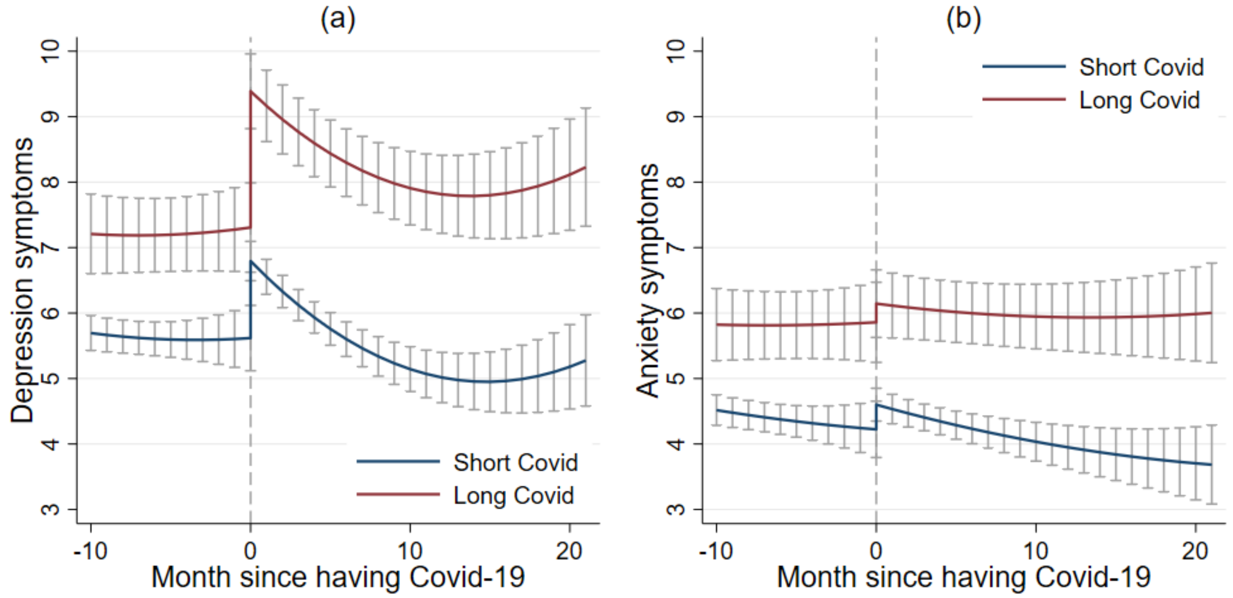


Fig S8. Predicted growth trajectories by Covid-19 group with 95% confidence intervals based on kernel matching *(NB these sensitivity analyses yielded less balanced matches so apparent differences in pre-infection mental health should be interpreted with caution)*

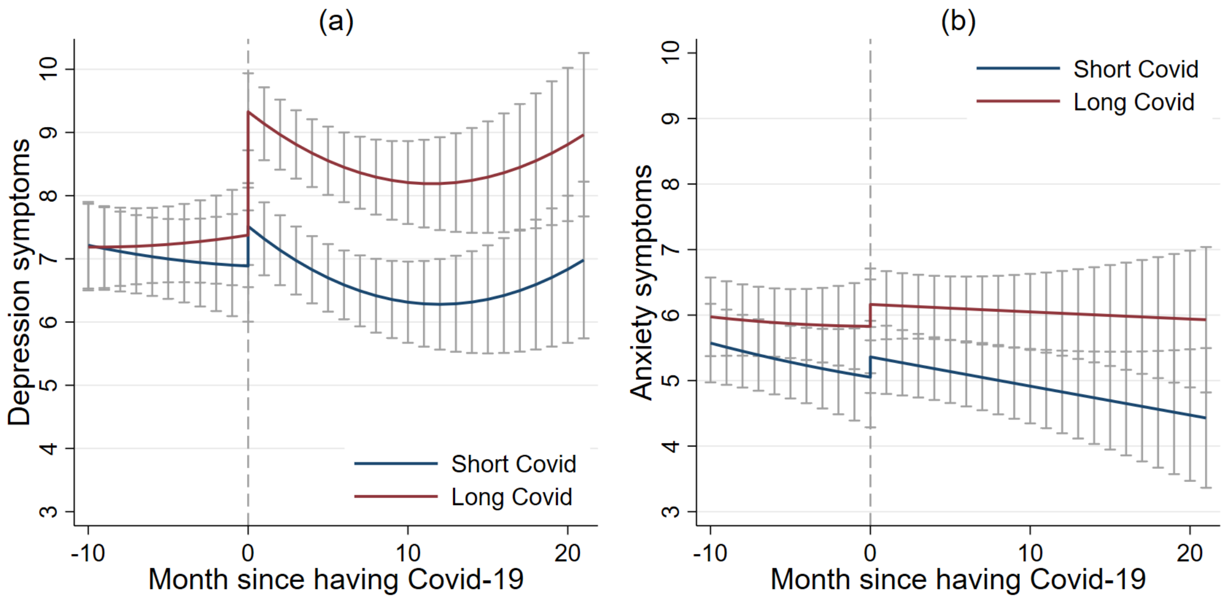


Fig S9. Predicted growth trajectories by Covid-19 group with 95% confidence intervals based on one-to-one matching excluding cases based on symptoms

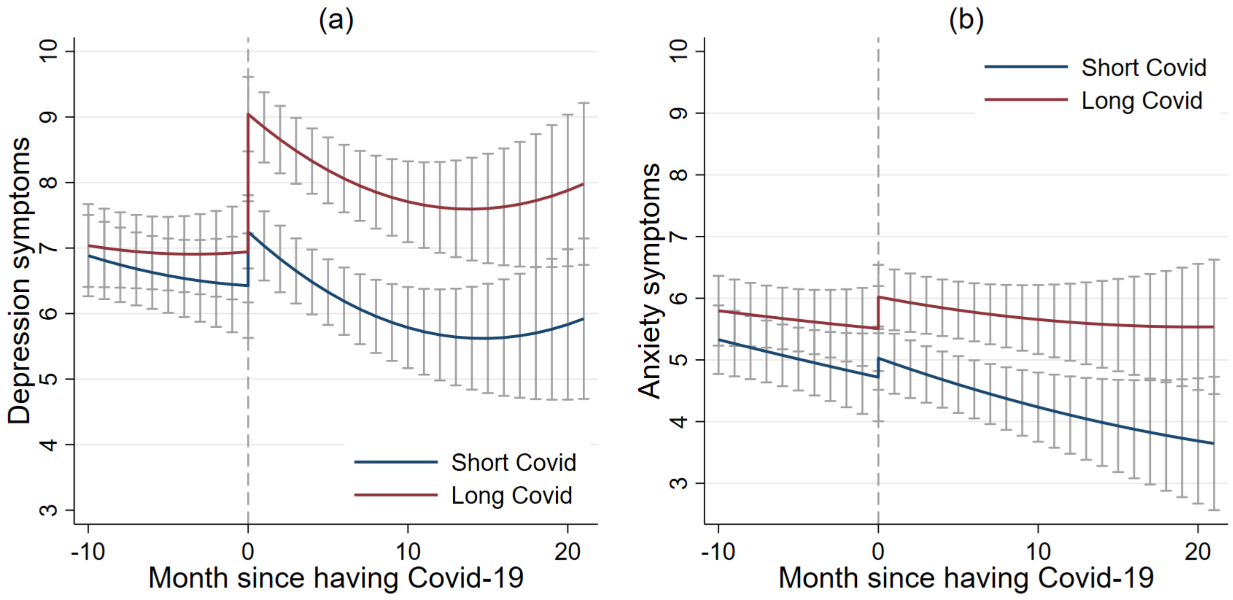


Fig S10. Predicted growth trajectories by Covid-19 group with 95% confidence intervals based on one-to-one matching excluding people who were unsure about their long Covid diagnosis

Table S1 Sample characteristics by matched Covid-19 group

| | | Long Covid (N=481) | Short Covid (N=481) |
|---|---|---|---|
| Gender | Men | 21.8% | 21.0% |
| | Women | 78.2% | 79.0% |
| Ethnicity | White | 94.2% | 95.0% |
| | Ethnic minority | 5.8% | 5.0% |
| Age group | 18-29 | 6.0% | 7.7% |
| | 30-45 | 31.6% | 30.8% |
| | 46-59 | 39.1% | 38.9% |
| | 60+ | 23.3% | 22.7% |
| Education | GCSE or below | 15.4% | 16.6% |
| | A levels or equivalent | 19.5% | 18.7% |
| | Degree or above | 65.1% | 64.7% |
| Annual income | <£16,000 | 14.3% | 14.8% |
| | ≤£16,000-29,999 | 23.7% | 22.7% |
| | ≤£30,000-59,999 | 38.3% | 41.0% |
| | ≤£60,000-89,999 | 16.2% | 15.0% |
| | ≥£90,000 | 7.5% | 6.7% |
| Employment status | Employed, keyworker | 39.1% | 40.1% |
| | Employed, non-keyworker | 32.6% | 31.0% |
| | Not employed | 28.3% | 28.9% |
| Area of living | City | 32.0% | 31.0% |
| | Town | 46.8% | 48.2% |
| | Rural | 21.2% | 20.8% |
| Living status | Living alone | 14.8% | 12.5% |
| | Living with adult only | 48.2% | 50.7% |
| | Living with children | 37.0% | 36.8% |
| Social contact | Twice a month or less | 35.3% | 33.7% |
| | Once or twice a week | 33.3% | 33.3% |
| | Three times a week or more | 31.4% | 33.1% |
| Number of close friends | | 4.7 (3.1) | 4.8 (3.1) |
| Physical health condition | Yes | 44.1% | 44.1% |
| | No | 55.9% | 55.9% |
| Mental health condition | Yes | 23.1% | 23.9% |
| | No | 76.9% | 76.1% |
| Baseline depressive symptoms | | 8.3 (5.9) | 8.2 (6.0) |
| Baseline anxiety symptoms | | 6.5 (5.4) | 6.4 (5.2) |
| Month had COVID-19 | | 10.5 (6.5) | 10.3 (7.1) |
| Confirmed by testing (COVID-19 or antibody test) | Yes | 74.6% | 72.8% |
| | No | 25.4% | 27.2% |
| Severe symptoms (hospitalised or had to stay in bed) | Yes | 81.7% | 82.5% |
| | No | 18.3% | 17.5% |

Table S2. Detailed Covid experience measures by matched Covid-19 group

| | | Long Covid | Short Covid | Total |
|---|---|---|---|---|
| Test | Yes, a medical professional has formally diagnosed me with long Covid | 112<br>23.3% | -- | 112<br>11.6% |
| | Yes, I have not been formally diagnosed but consider myself to have Long Covid | 369<br>76.7% | -- | 369<br>38.4% |
| | No, I do not consider myself to have Long Covid | -- | 369<br>76.7% | 369<br>38.4% |
| | I am unsure | -- | 112<br>23.3% | 112<br>11.6% |
| | **Total** | **481**<br>**100%** | **481**<br>**100%** | **962**<br>**100%** |
| Severity (first 1-2 weeks) | I was hospitalised | 30<br>6.2% | 16<br>3.3% | 46<br>4.8% |
| | I experienced symptoms and had to rest in bed | 363<br>75.5% | 381<br>79.2% | 744<br>77.3% |
| | I experienced symptoms but was able to carry on with daily activities | 79<br>16.4% | 71<br>14.8% | 150<br>15.6% |
| | I was asymptomatic | 9<br>1.9% | 13<br>2.7% | 22<br>2.3% |
| | **Total** | **481**<br>**100%** | **481**<br>**100%** | **962**<br>**100%** |
| Symptoms | My symptoms were worse at the beginning (the first 1-2 weeks) and then got better | 23<br>4.8% | 179<br>37.6% | 202<br>21.2% |
| | My symptoms were worse at the beginning (the first 1-2 weeks) and then mostly got better but some lingered | 152<br>32.0% | 167<br>35.1% | 319<br>33.5% |
| | After the first 1-2 weeks, my symptoms got better but then the same symptoms kept coming back | 67<br>14.1% | 21<br>4.4% | 88<br>9.3% |
| | After the first 1-2 weeks, my symptoms got better but I then developed new symptoms | 60<br>12.6% | 15<br>3.2% | 75<br>7.9% |
| | Most of my symptoms lasted for 2-3 weeks | 25<br>5.3% | 39<br>8.2% | 64<br>6.7% |
| | Most of my symptoms lasted for 4-12 weeks | 62<br>13.1% | 23<br>4.8% | 85<br>8.9% |
| | Most of my symptoms lasted for more than 12 weeks | 81<br>17.1% | 6<br>1.3% | 87<br>9.2% |
| | I cannot answer this question (e.g. you had COVID very recently) | 5<br>1.1% | 26<br>5.5% | 31<br>3.3% |
| | **Total** | **475**<br>**100%** | **476**<br>**100%** | **951**<br>**100%** |
